# Age-related Muscle Fat Infiltration in Lung Screening Participants: Impact of Smoking Cessation

**DOI:** 10.1101/2023.12.05.23299258

**Authors:** Kaiwen Xu, Thomas Z. Li, James G. Terry, Aravind R. Krishnan, Stephen A. Deppen, Yuankai Huo, Fabien Maldonado, J. Jeffrey Carr, Bennett A. Landman, Kim L. Sandler

**Affiliations:** Department of Computer Science, Vanderbilt University, Nashville, Tennessee; Department of Biomedical Engineering, Vanderbilt University, Nashville, Tennessee; School of Medicine, Vanderbilt University, Nashville, Tennessee; Department of Electrical and Computer Engineering, Vanderbilt University, Nashville, Tennessee; Department of Radiology, Vanderbilt University Medical Center, Nashville, Tennessee; Department of Thoracic Surgery, Vanderbilt University Medical Center, Nashville, Tennessee; Department of Medicine, Vanderbilt University Medical Center, Nashville, Tennessee; Department of Biomedical Informatics, Vanderbilt University Medical Center, Nashville, Tennessee

**Keywords:** skeletal muscle, thorax, computed tomography, artificial

## Abstract

**Rationale:** Skeletal muscle fat infiltration progresses with aging and is worsened among individuals with a history of cigarette smoking. Many negative impacts of smoking on muscles are likely reversible with smoking cessation.

**Objectives:** To determine if the progression of skeletal muscle fat infiltration with aging is altered by smoking cessation among lung cancer screening participants.

**Methods:** This was a secondary analysis based on the National Lung Screening Trial. Skeletal muscle attenuation in Hounsfield unit (HU) was derived from the baseline and follow-up low-dose CT scans using a previously validated artificial intelligence algorithm. Lower attenuation indicates greater fatty infiltration. Linear mixed-effects models were constructed to evaluate the associations between smoking status and the muscle attenuation trajectory.

**Measurements and Main Results:** Of 19,019 included participants (age: 61 years, 5 [SD]; 11,290 males), 8,971 (47.2%) were actively smoking cigarettes. Accounting for body mass index, pack-years, percent emphysema, and other confounding factors, actively smoking predicted a lower attenuation in both males (β_0_ =-0.88 HU, *P*<.001) and females (β_0_ =−0.69 HU, *P*<.001), and an accelerated muscle attenuation decline-rate in males (β_l_=−0.08 HU/y, *P*<.05). Age-stratified analyses indicated that the accelerated muscle attenuation decline associated with smoking likely occurred at younger age, especially in females.

**Conclusions:** Among lung cancer screening participants, active cigarette smoking was associated with greater skeletal muscle fat infiltration in both males and females, and accelerated muscle adipose accumulation rate in males. These findings support the important role of smoking cessation in preserving muscle health.

## 1. Introduction

Skeletal muscle dysfunction is a primary extrapulmonary manifestation of chronic obstructive pulmonary disease (COPD), a condition highly associated with chronic cigarette smoking (1–5). While some of the adverse effects on the muscles can be attributed to the decline in lung function, emerging evidence from both small clinical studies and animal models indicates that smoking *per se* can alter the muscle anatomy and impair muscle function independently of pulmonary disease status (4, 6–8). In distinction to the damage to lung parenchyma, many negative impacts of smoking on muscles are likely reversible with smoking cessation (4, 8). U.S. Preventive Services Task Force recommends lung cancer screening for high risk individuals, characterized by a significant tobacco use history and advanced age (9), which also forms a population subject to declining physiological reserve and at elevated risk for developing frailty (10–12). Furthermore, skeletal muscle fat infiltration progresses with aging and is associated with reduced muscle strength at older age (13–15). Excessive fatty infiltration in non-adiposity issues is closely linked to insulin resistance, with cigarette smoking identified as significant contributing factor (13, 16–18). Promoting smoking cessation is one important component in lung cancer screening (19–21). Nevertheless, the potential benefits of cessation in preserving muscle health have not been well characterized within the screening population.

Muscle attenuation (or radiodensity), as determined by computed tomography (CT), offers a reliable and noninvasive characterization for fatty infiltration in muscles (22, 23). Previous studies have reported significant health implications of muscle attenuation, including incident mobility limitations at older age (24, 25); lung function trajectory in asthma (26); and mortality risks in asymptomatic cancer screening populations (27–30). Moreover, increasing evidence support fatty infiltration as a more consistent signature of worsening muscle health compared to the traditional sarcopenia characterized by reduced muscle mass (14, 17, 25, 27).

The associations between cigarette smoking and increased skeletal muscle fat infiltration have been confirmed in previous observational studies (31, 32). These studies were conducted in cross-sectional settings, with a primary focus on evaluating the influence of smoking on muscle adiposity when compared to non-smoking healthy controls. Our primary focus was to evaluate the potential benefits of smoking cessation in decelerating fatty infiltration among individuals with a heavy smoking history. By utilizing the lung screening non-contrast low-dose CT (LDCT) and a previously validated artificial intelligence algorithm (27, 33, 34), we obtained the longitudinal muscle attenuation data for a population-scale lung cancer screening cohort. We hypothesized that, in addition to lowered muscle attenuation, active smoking is associated with a more rapid muscle attenuation decline rate, signifying an accelerated progression of fatty infiltration associated with ongoing smoking. Furthermore, we investigated the correlation between smoking cessation duration with the trajectory of muscle attenuation among those who had quit smoking.

## 2. Methods

This observational study was a secondary analysis of the National Lung Screening Trial (NLST, *ClinicalTrials.gov number: NCT00047385*), a multicenter randomized controlled trial to comparing chest LDCT with radiography for lung cancer screening (35, 36). The details of NLST protocols have been reported in previous publications (35, 36). Briefly, the inclusion criteria selected those aged 55–74 years and with a history of smoking of at least 30 pack-years and had smoked within the past 15 years. Participant enrollment was conducted from August 2002 through April 2004. The trial was approved by the institutional review board at each participating medical institution. Written informed consent was provided by each participant. The authors obtained permission to access and analyze the de-identified data collected by NLST researchers.

### 2.1 Cohort Selection and Data

NLST participants were invited to undergo a baseline and two annual follow-up screenings. Those randomized to the CT arm underwent LDCT imaging, following standard manufacture-specific image acquisition protocols as outlined in web-distributed technique charts (37, 38). In this study, we only included participants who completed all three screenings. We exclusively included images with slice spacing smaller than 2.5 mm, total scan length lager than 200 mm, and with reconstruction kernels among the manufacture-specific soft tissue kernels listed by NLST protocol mandated by American College of Radiology Imaging Network (37). Participants without a valid baseline LDCT were excluded from analysis. Additionally, we excluded the follow-up LDCT scans conducted using a different scanner manufacture or model than the one utilized for baseline LDCT scans. This exclusion aimed to mitigate the potential influence of changing scanner platform on the longitudinal trajectory of CT-derived measurements.

### 2.2 Clinical Characteristics

The demographic characteristics, smoking behavior, and disease history of each NLST participant were self-reported through a questionnaire completed at the time of randomization to the two arms of the trial. We extracted characteristics known or potentially associated with skeletal muscle fat infiltration. These included age, race, BMI (kg/m^2^); education level, marital status, current smoking status, pack-year, smoking quit time (reported in years for those who had quit smoking), diagnostic history of cardiovascular diseases, diabetes, and COPD. Participants with incomplete clinical data elements were excluded from statistical analysis. The specific procedures for cohort selection are outlined in Figure 1.

**Figure 1:**
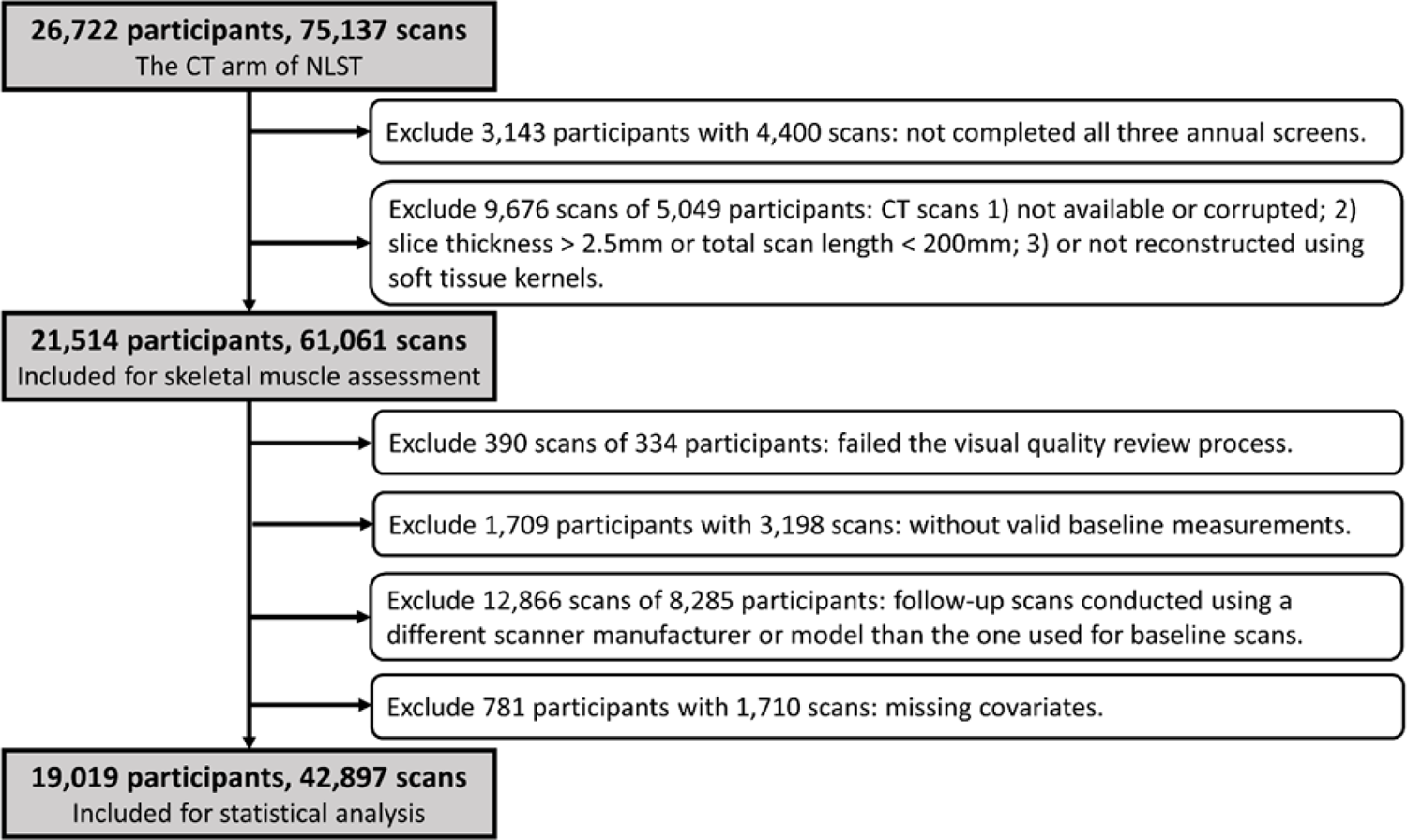
Cohort selection flowchart. NLST=National Lung Screening Trial.

### 2.3 Quantitative CT

Muscle attenuation was derived from the baseline and follow-up LDCT scans using a previously validated AI algorithm. The software is publicly available in the form of a Docker container image (39) maintained at Docker Hub™. The user instructions are documented at https://github.com/MASILab/S-EFOV. The development and evaluation of the algorithm have been described in previous publications (27, 33, 34). An example measurement is presented in Figure 2.

**Figure 2:**
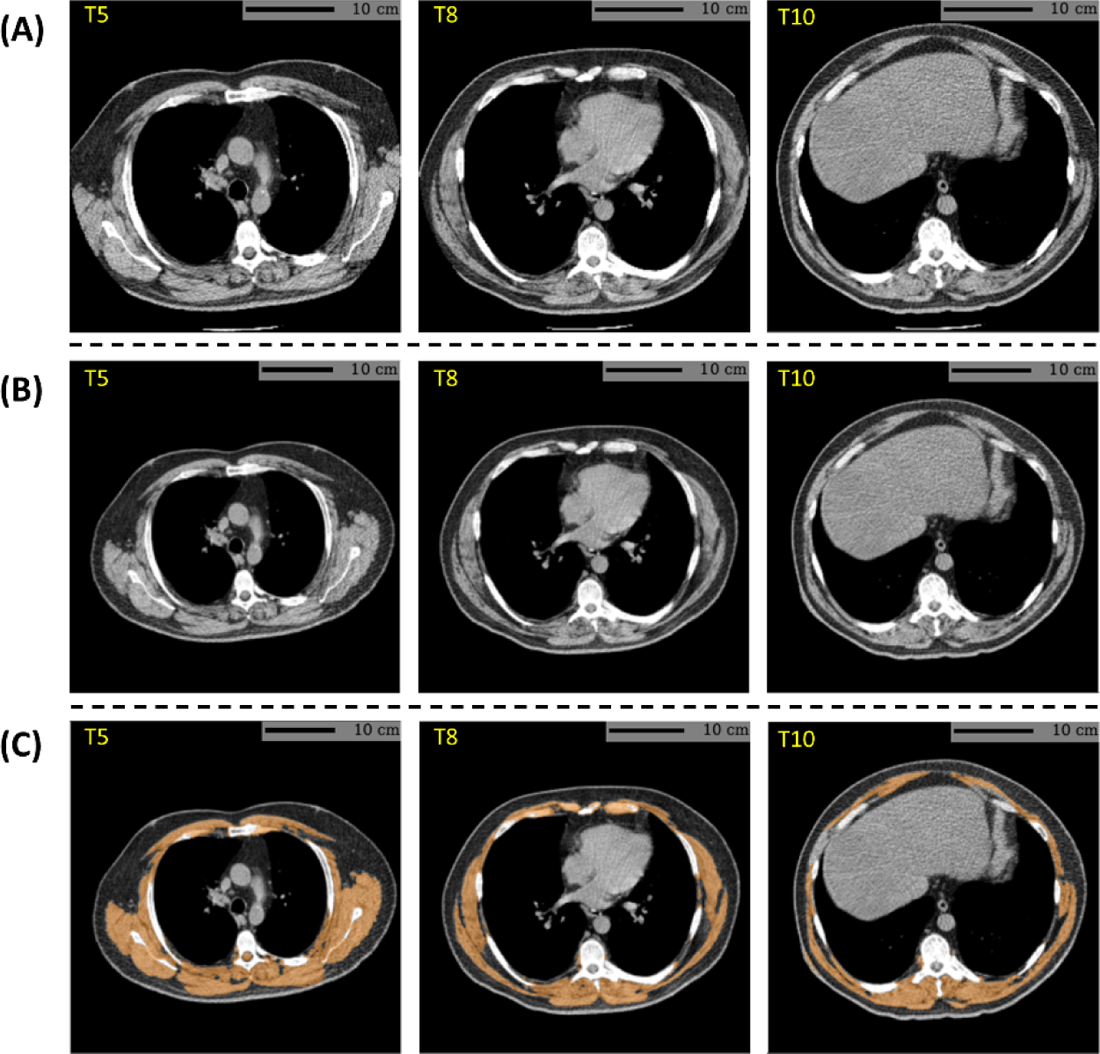
Example skeletal muscle assessment using lung cancer screening low-dose CT scan of a male participant with a BMI of 35.1 kg/m and a smoking history of 41 years (61.5 pack-years). (A) CT axial plane levels corresponding the mid-location of the fifth (T5), eighth (T8), and 10th (T10) vertebral bodies were predicted. (B) The field-of-view of each CT section with body section truncation was extended with missing body section imputation. (C) Cross-sectional regions corresponding to the skeletal muscle were delineated on the field-of-view extended sections. Skeletal muscle attenuation was defined as the averaged radiodensity of muscle regions across the three levels. In this case, the algorithm reported a skeletal muscle attenuation of 17.5 HU, which fell within the lowest quartile (< 27.6 HU) among male participants. BMI=body mass index. HU=Hounsfield unit.

To address the potential influence of lung function impairment on muscle adiposity, we incorporated percent emphysema, determined for each included LDCT through a fully automatic procedure. The lung regions in a LDCT volume were identified using a publicly available deep learning lung segmentation algorithm (40). Percent emphysema was defined as the percentage of lung volume containing voxels with values less than −950 HU, following established practice (41, 42).

Since all image assessments were performed by automatic algorithms, a secondary independent analysis was not conducted. However, the quality of the assessments for all included LDCT images underwent visual review. Screenings with failed assessments were subsequently excluded from the statistical analysis (Figure 1).

### 2.4 Statistical Analysis

Descriptive statistics are presented as mean (SD) for continuous variables and as a percentage for categorical variables. Annualized change rates were estimated from available longitudinal measurements using linear regression. Differences between current and former tobacco users were separately assessed for males and females. Mann-Whitney U tests were employed to compare continuous variables, while chi-square tests were used for categorical variables.

The central aim of this study was to investigate the association between smoking status (current vs. former) and the progression of skeletal muscle fat infiltration, assessed by muscle attenuation, over time. Additionally, we examined the influence of the time since cessation on muscle attenuation trajectory among individuals who had quit smoking. Linear mixed-effects models adjusted for confounding factors were employed to investigate these potential associations. These models were constructed to predict muscle attenuation at each measurement time, accounting for intra-participant correlation through the inclusion of random intercepts and slopes for each participant. Furthermore, the models allowed for the inclusion of participants with incomplete follow-up data (43). The primary variables of interest were included through a main effect term, as well as a linear trend term in form of an intersection with screening time in years since baseline. After fitting the model to the longitudinal muscle attenuation data, the regression coefficient of the main effect term (denoted asβ_0_) signifies the cross-sectional association between the variable of interest and muscle attenuation. Meanwhile, the regression coefficient of the linear trend term (denoted asβ_l_) reflects the variable’s influence on the annualized muscle attenuation change rate.

A series of models were fit by incrementally adding confounding factors: Model 1 – unadjusted; Model 2 – adjusted for demographic and social economic factors, including age, race/ethnicity, education level, and marital status; Model 3 – adjusted additionally history of comorbidities, including cardiovascular diseases, diabetes, and COPD; Model 4 – adjusted additionally for BMI; Model 5 – adjusted additionally for pack-years; Model 6 – adjusted additionally for percent emphysema. All confounding factors, except percent emphysema, were included via a main effect term and a linear trend term using values obtained at baseline. Percent emphysema was incorporated as a transient effect (or time-varying) term (44, 45).

Analyses stratified by BMI and age were performed to assess their influence on the associations between smoking and muscle attenuation trajectory. Regarding BMI, the cohort was categorized into normal/underweight (BMI < 25), overweight (25 ≤ BMI < 30), and obese (BMI≥30) groups. In terms of age, the cohort was divided into a younger group (aged between 55 and 64 years) and an older group (aged between 65 and 74 years).

All analyses were stratified by sex. The regression coefficients (bothβ_0_ andβ_l_) for factors of interest and their associated *P* values are reported. We *a priori* defined *P* values less than 0.05 as indictive of statistically significant. All statistical tests were two-sided. Statistical analyses were conducted using R 4.1.2 (R Foundation for Statistical Computing).

## 3. Results

### 3.1 Cohort Characteristics

A total of 19,019 participants with 42,897 LDCT screening scans were included in the statistical analysis. The breakdown of excluded participants and LDCT scans for each criterion is detailed in Figure 1. Among those included, the mean age was 61 years (standard deviation [SD], 5 years); 7,729 (40.6%) were female; and 8,971 (47.2%) were actively smoking cigarette at baseline. Among those who had quit smoking, the mean duration since cessation was 7.3 years (SD, 4.7 years). A total of 13,970 participants (73.5%) had at least one valid follow-up scan, with 12,647 (66.5%) having a valid year-1 follow-up scan and 11,318 (59.5%) having a valid year-2 follow-up scan, allowing for estimation (linear regression) of the annualized change rate of percent emphysema and muscle attenuation (Table 1). When not adjusted for other factors, active smoking was linked to an accelerated decline rate of muscle attenuation, while those who had quit smoking exhibited a lower baseline attenuation value. Moreover, older age, higher education level, a marital status of “married or living as married,” an increased diagnosis history of cardiovascular disease or diabetes, and more severe emphysema were observed among those who had quit smoking. Figure 3 presents the muscle attenuation data stratified by age and BMI, revealing a negative association between muscle attenuation and BMI as well as age. Consistent declines of muscle attenuation with aging were observed across all subgroups.

**Figure 3:**
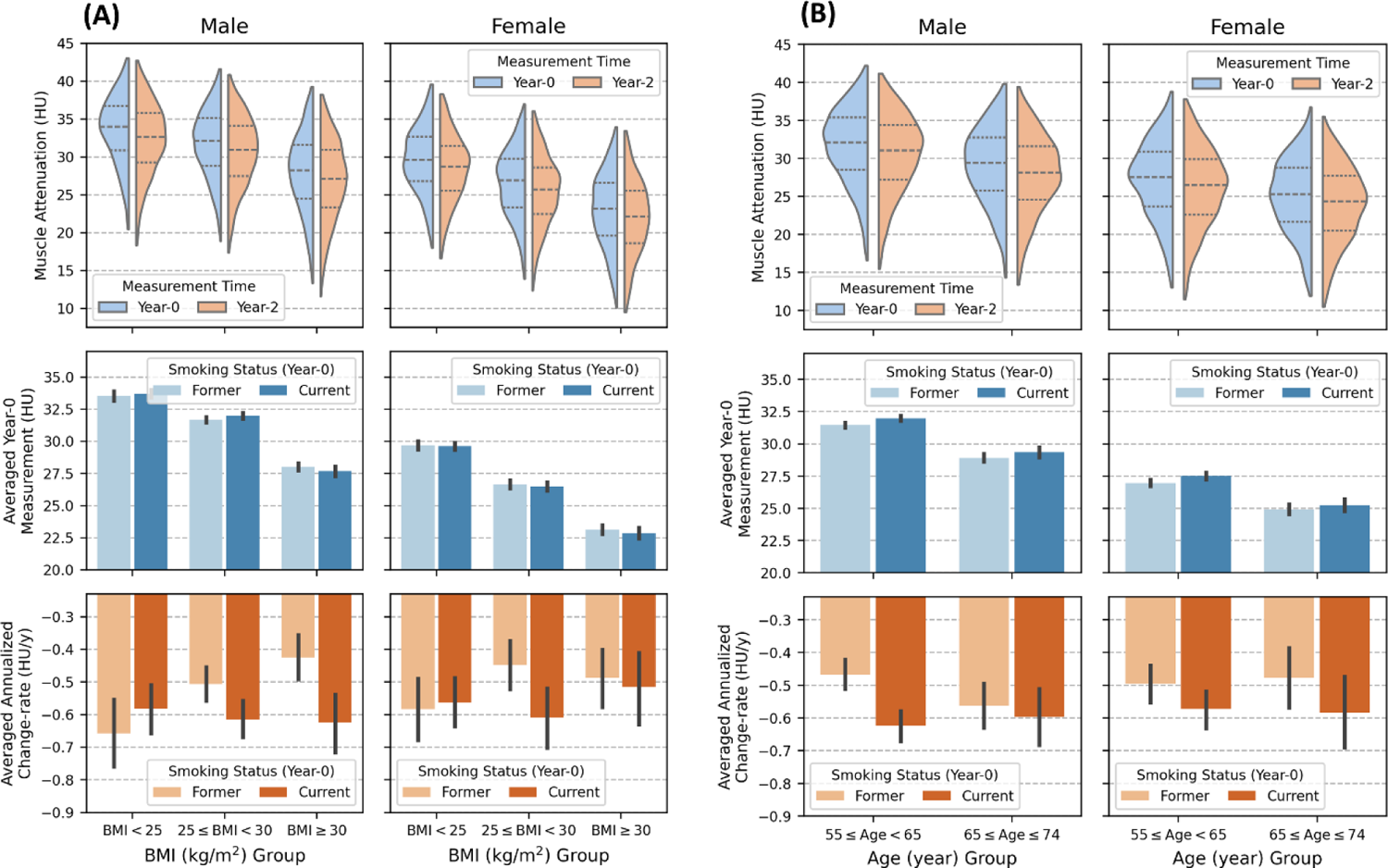
Sex-stratified muscle attenuation data, further stratified by age and body mass index (BMI), ar presented. (A) illustrates data stratified by body mass index (BMI), while (B) shows data stratified by age. In both (A) and (B), the left columns show subplots for males, and the right columns show subplots for females. The to rows illustrate the distribution of attenuation measurements at baseline (year-0) screening (blue) and measurements at year-2 screening (orange) for those with valid year-2 follow-up scan. The middle rows display th estimated mean baseline attenuation for each subgroup, stratified by baseline smoking status. The bottom rows show the estimated mean annualized attenuation change-rage (HU/y) for each subgroup, stratified by baselin smoking status. HU= Hounsfield Unit. BMI=Body Mass Index. COPD=Chronic Obstructive Pulmonary Disease.

**Table 1:**
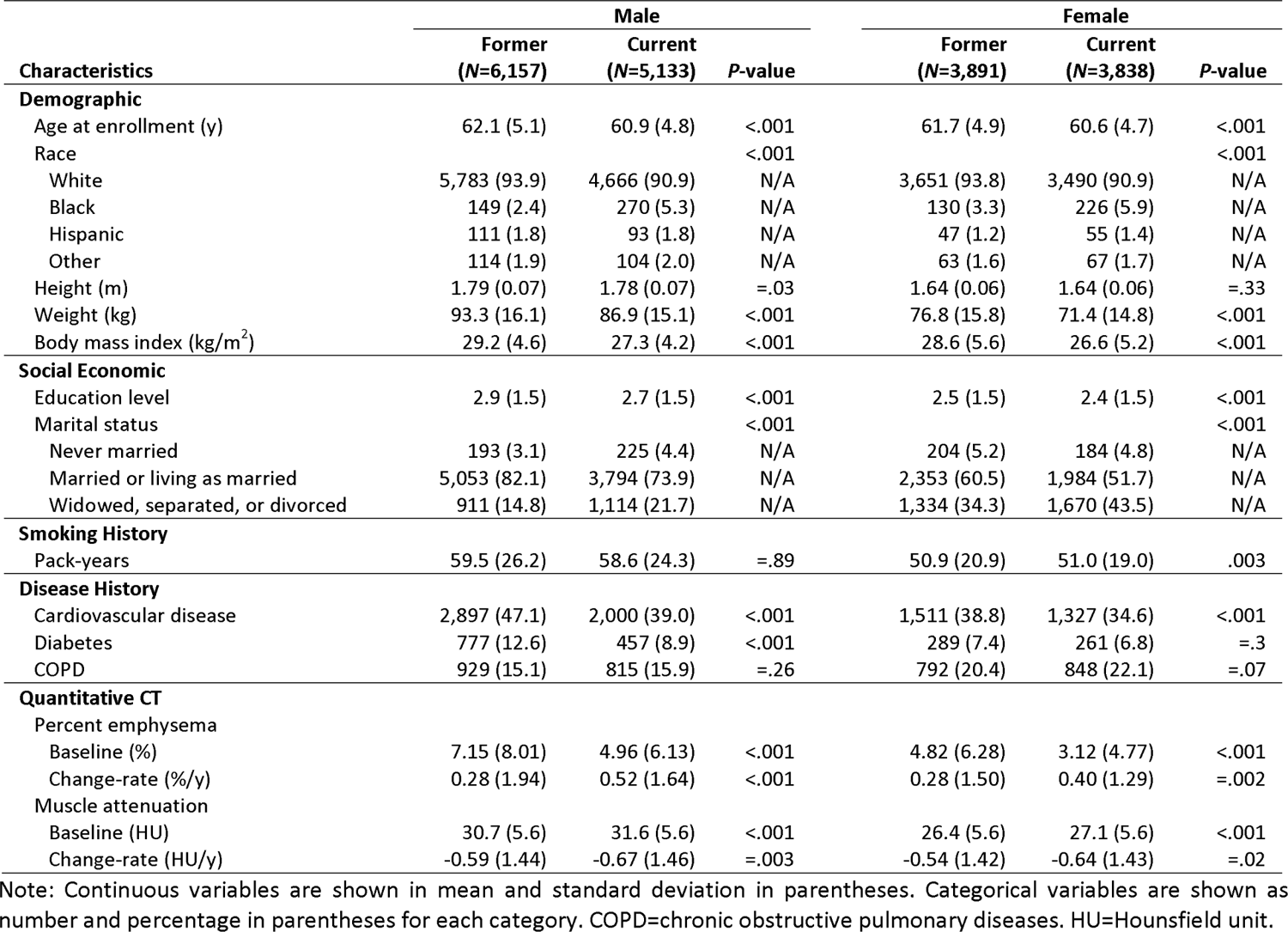
Cohort characteristics.

| Characteristics | Male |  |  | Female |  |  |
| --- | --- | --- | --- | --- | --- | --- |
|  | Former<br>(N=6,157) | Current<br>(N=5,133) | P-value | Former<br>(N=3,891) | Current<br>(N=3,838) | P-value |
| **Demographic** |  |  |  |  |  |  |
| Age at enrollment (y) | 62.1 (5.1) | 60.9 (4.8) | <.001 | 61.7 (4.9) | 60.6 (4.7) | <.001 |
| Race |  |  | <.001 |  |  | <.001 |
| White | 5,783 (93.9) | 4,666 (90.9) | N/A | 3,651 (93.8) | 3,490 (90.9) | N/A |
| Black | 149 (2.4) | 270 (5.3) | N/A | 130 (3.3) | 226 (5.9) | N/A |
| Hispanic | 111 (1.8) | 93 (1.8) | N/A | 47 (1.2) | 55 (1.4) | N/A |
| Other | 114 (1.9) | 104 (2.0) | N/A | 63 (1.6) | 67 (1.7) | N/A |
| Height (m) | 1.79 (0.07) | 1.78 (0.07) | =.03 | 1.64 (0.06) | 1.64 (0.06) | =.33 |
| Weight (kg) | 93.3 (16.1) | 86.9 (15.1) | <.001 | 76.8 (15.8) | 71.4 (14.8) | <.001 |
| Body mass index (kg/m<sup>2</sup>) | 29.2 (4.6) | 27.3 (4.2) | <.001 | 28.6 (5.6) | 26.6 (5.2) | <.001 |
| **Social Economic** |  |  |  |  |  |  |
| Education level | 2.9 (1.5) | 2.7 (1.5) | <.001 | 2.5 (1.5) | 2.4 (1.5) | <.001 |
| Marital status |  |  | <.001 |  |  | <.001 |
| Never married | 193 (3.1) | 225 (4.4) | N/A | 204 (5.2) | 184 (4.8) | N/A |
| Married or living as married | 5,053 (82.1) | 3,794 (73.9) | N/A | 2,353 (60.5) | 1,984 (51.7) | N/A |
| Widowed, separated, or divorced | 911 (14.8) | 1,114 (21.7) | N/A | 1,334 (34.3) | 1,670 (43.5) | N/A |
| **Smoking History** |  |  |  |  |  |  |
| Pack-years | 59.5 (26.2) | 58.6 (24.3) | =.89 | 50.9 (20.9) | 51.0 (19.0) | .003 |
| **Disease History** |  |  |  |  |  |  |
| Cardiovascular disease | 2,897 (47.1) | 2,000 (39.0) | <.001 | 1,511 (38.8) | 1,327 (34.6) | <.001 |
| Diabetes | 777 (12.6) | 457 (8.9) | <.001 | 289 (7.4) | 261 (6.8) | =.3 |
| COPD | 929 (15.1) | 815 (15.9) | =.26 | 792 (20.4) | 848 (22.1) | =.07 |
| **Quantitative CT** |  |  |  |  |  |  |
| Percent emphysema |  |  |  |  |  |  |
| Baseline (%) | 7.15 (8.01) | 4.96 (6.13) | <.001 | 4.82 (6.28) | 3.12 (4.77) | <.001 |
| Change-rate (%/y) | 0.28 (1.94) | 0.52 (1.64) | <.001 | 0.28 (1.50) | 0.40 (1.29) | =.002 |
| Muscle attenuation |  |  |  |  |  |  |
| Baseline (HU) | 30.7 (5.6) | 31.6 (5.6) | <.001 | 26.4 (5.6) | 27.1 (5.6) | <.001 |
| Change-rate (HU/y) | -0.59 (1.44) | -0.67 (1.46) | =.003 | -0.54 (1.42) | -0.64 (1.43) | =.02 |

Note: Continuous variables are shown in mean and standard deviation in parentheses. Categorical variables are shown as number and percentage in parentheses for each category. COPD=chronic obstructive pulmonary diseases. HU=Hounsfield unit.

### 3.2 Longitudinal Trajectory of Muscle Attenuation

The fitted regression coefficients for the primary variables of interest are presented in Table 2. In fully adjusted models, actively smoking was associated with a reduced baseline muscle attenuation in both males (=−0.882 HU, [SE: 0.091]; *P*<.001) and females (=−0.688 HU, [SE: 0.110]; *P*<.001). The decline in attenuation was more rapid in males who continued to smoke than in males who had quit smoking (=−0.079 HU/y, [SE: 0.028]; *P*=.005), but this trend was no longer significant in females (=−0.045 HU/y, [SE: 0.035]; *P*=.2). Among those who had quit smoking, the number of years since cessation was directly associated with an increased baseline attenuation in both males (β_0_=0.045 HU/y, [SE: 0.012]; *P*<.001) and females (β_0_=0.035 HU/y, [SE: 0.016]; *P*=.03). However, there was no association observed between duration since cessation and longitudinal change rate of attenuation in either males or females. Results of intermediate models with incrementally added confounding factors are also presented in Table 2. Notably, the fitted main effect coefficients of both current smoking status and smoking quit time changed dramatically after the inclusion of BMI as confounding factor.

**Table 2:** Incremental Models for Assessing Association of Baseline Smoking Status with Longitudinal Trajectory of Muscle Attenuation.

| Primary predictor, model | Male |  | Female |  |
| --- | --- | --- | --- | --- |
|  | Main effect | Linear trend | Main effect | Liner trend |
| Smoking status (current vs. former) |  |  |  |  |
| Model 1 | 0.879 (0.106) [$<.001$] | -0.108 (0.028) [$<.001$] | 0.721 (0.128) [$<.001$] | -0.077 (0.034) [= .02] |
| Model 2 | 0.544 (0.103) [$<.001$] | -0.105 (0.028) [$<.001$] | 0.493 (0.127) [$<.001$] | -0.069 (0.034) [= .043] |
| Model 3 | 0.329 (0.101) [$<.001$] | -0.114 (0.028) [$<.001$] | 0.416 (0.123) [$<.001$] | -0.069 (0.034) [= .043] |
| Model 4 | -0.695 (0.091) [$<.001$] | -0.094 (0.029) [= .001] | -0.622 (0.110) [$<.001$] | -0.051 (0.035) [= .15] |
| Model 5 | -0.681 (0.090) [$<.001$] | -0.094 (0.029) [= .002] | -0.607 (0.109) [$<.001$] | -0.050 (0.035) [= .16] |
| Model 6 | -0.882 (0.091) [$<.001$] | -0.079 (0.028) [= .005] | -0.688 (0.110) [$<.001$] | -0.045 (0.035) [= .2] |
| †Smoking quit time (y) |  |  |  |  |
| Model 1 | -0.023 (0.015) [= .12] | 0.001 (0.004) [= .89] | -0.038 (0.019) [= .051] | 0.003 (0.005) [= .62] |
| Model 2 | 0.029 (0.015) [= .051] | 0.002 (0.004) [= .57] | 0.005 (0.019) [= .78] | 0.002 (0.005) [= .7] |
| Model 3 | 0.026 (0.014) [= .07] | 0.003 (0.004) [= .51] | 0.011 (0.019) [= .56] | 0.002 (0.005) [= .74] |
| Model 4 | 0.051 (0.012) [$<.001$] | 0.002 (0.004) [= .64] | 0.040 (0.016) [= .01] | 0.001 (0.005) [= .88] |
| Model 5 | 0.044 (0.012) [$<.001$] | 0.003 (0.004) [= .52] | 0.034 (0.016) [= .04] | 0.001 (0.005) [= .87] |
| Model 6 | 0.045 (0.012) [$<.001$] | 0.002 (0.004) [= .63] | 0.035 (0.016) [= .03] | 0.000 (0.005) [= .97] |

Note: Displayed numbers are estimated regression coefficients, standard errors in parentheses, and $P$ values in brackets. The main effect terms assess the association with baseline muscle attenuation, while the linear trend terms (by interaction with time) assess the association with linear longitudinal change rate of muscle attenuation. †Analyses with smoking quit time as primary predictor were performed among those who had quit smoking at baseline.
Model 1: Unadjusted
Model 2: Adjusted for age, race, education level, and marital status
Model 3: Model 2 + Diagnostic history of cardiovascular diseases, diabetes, and COPD
Model 4: Model 3 + Body mass index
Model 5: Model 4 + Pack-year
Model 6: Model 5 + Percent emphysema

In BMI-stratified analyses (Table 3), a higher BMI was associated with decreased baseline muscle attenuation, while the overall longitudinal attenuation decline rate was more rapid in leaner individuals. Associations of active smoking with lowered baseline attenuation were observed in all BMI subgroups, and the magnitude of these associations increased with level of obesity. The most substantial impact of active smoking on attenuation change rate was observed in the obese group (β_l_=−0.120 HU/y, [SE: 0.054]) in males. However, in females, the most substantial impact was observed in overweight group (β_l_=−0.140 HU/y, [SE: 0.056]) instead of obese group. In both males and females, the change rate of attenuation was not altered by smoking status in the normal/underweight groups.

**Table 3:** Association of baseline smoking status with longitudinal trajectory of muscle attenuation in analysis stratified by body mass index. The first part shows the characteristics of each subgroup, while the second part shows regression coefficients of fully adjusted linear mix-effects models for each subgroup.

|  | Male, BMI (kg/m<sup>2</sup>) Group |  |  | Female, BMI (kg/m<sup>2</sup>) Group |  |  |
| --- | --- | --- | --- | --- | --- | --- |
|  | BMI<25 | 25≤BMI<30 | BMI≥30 | BMI<25 | 25≤BMI<30 | BMI≥30 |
| **Cohort characteristics** |  |  |  |  |  |  |
| No. participants | 2,484 | 5,422 | 3,384 | 2,805 | 2,747 | 2,177 |
| Age (y), SD | 61.9 (5.2) | 61.7 (5.1) | 61.2 (4.8) | 61.1 (4.9) | 61.3 (4.9) | 60.9 (4.7) |
| Smoking status, % |  |  |  |  |  |  |
| Current | 1,548 (62.3) | 2,435 (44.9) | 1,150 (34.0) | 1,677 (59.8) | 1,304 (47.5) | 857 (39.4) |
| Former | 936 (37.7) | 2,987 (55.1) | 2,234 (66.0) | 1,128 (40.2) | 1,443 (52.5) | 1,320 (60.6) |
| Disease history, % |  |  |  |  |  |  |
| CVD | 774 (31.2) | 2,249 (41.5) | 1,874 (55.4) | 767 (27.3) | 997 (36.3) | 1,074 (49.3) |
| Diabetes | 131 (5.3) | 447 (8.8) | 626 (18.5) | 67 (2.4) | 155 (5.6) | 328 (15.1) |
| COPD | 490 (19.7) | 713 (13.2) | 541 (16.0) | 571 (20.4) | 567 (20.6) | 502 (23.1) |
| Muscle Attenuation |  |  |  |  |  |  |
| Year-0 (HU), SD | 33.7 (5.0) | 32.0 (4.9) | 27.9 (5.6) | 29.6 (4.7) | 26.7 (5.0) | 23.0 (5.5) |
| Change rate (HU/y), SD | -0.68 (1.52) | -0.62 (1.41) | -0.60 (1.48) | -0.63 (1.39) | -0.60 (1.43) | -0.52 (1.46) |
| **Association of smoking status (current vs. former) with muscle attenuation trajectory** |  |  |  |  |  |  |
| Main effect |  |  |  |  |  |  |
| Coefficient, SE | -0.528 (0.196) | -0.692 (0.125) | -1.299 (0.179) | -0.470 (0.175) | -0.722 (0.181) | -0.976 (0.224) |
| P Value | =.005 | <.001 | <.001 | =.007 | <.001 | <.001 |
| Change-rate |  |  |  |  |  |  |
| Coefficient, SE | 0.001 (0.063) | -0.097 (0.039) | -0.120 (0.054) | 0.028 (0.059) | -0.140 (0.056) | -0.025 (0.070) |
| P Value | =.98 | =.01 | =.03 | =.64 | =.01 | =.72 |
| **†Association of smoking quit time (y) with muscle attenuation trajectory** |  |  |  |  |  |  |
| Main effect |  |  |  |  |  |  |
| Coefficient, SE | 0.021 (0.031) | 0.058 (0.017) | 0.031 (0.022) | 0.063 (0.028) | 0.007 (0.026) | 0.035 (0.031) |
| P Value | =.50 | <.001 | =.16 | =.02 | =.78 | =.25 |
| Change-rate |  |  |  |  |  |  |
| Coefficient, SE | 0.008 (0.010) | 0.000 (0.006) | 0.002 (0.007) | -0.012 (0.010) | 0.009 (0.008) | 0.002 (0.010) |
| P Value | =.40 | =.96 | =.73 | =.20 | =.26 | =.85 |

†Analyses with smoking quit time as primary predictor were performed among those who had quit smoking at baseline.
CVD=cardiovascular disease. COPD=chronic obstructive pulmonary diseases

In age-stratified analyses (Table 4), older age was associated with lower baseline muscle attenuation. The overall attenuation change rate was a relatively consistent across both age groups in both males and females. Active smoking-associated acceleration in attenuation decline rate was only observed in younger males (β_l_=−0.120 HU/y, [SE: 0.033]), whereas associations of active smoking with decreased baseline attenuation were observed in all age subgroups in both males and females. Furthermore, a longer duration since smoking cessation correlated with increased baseline attenuation in both younger (β_0_=0.042 HU/y, [SE: 0.015]) and older (β_0_=0.054 HU/y, [SE: 0.022]) groups in males. In younger females, a similar association was observed (β_0_=0.058 HU/y, [SE: 0.019]), but not in older females (β_0_=−0.019 HU/y, [SE: 0.031]).

**Table 4:**
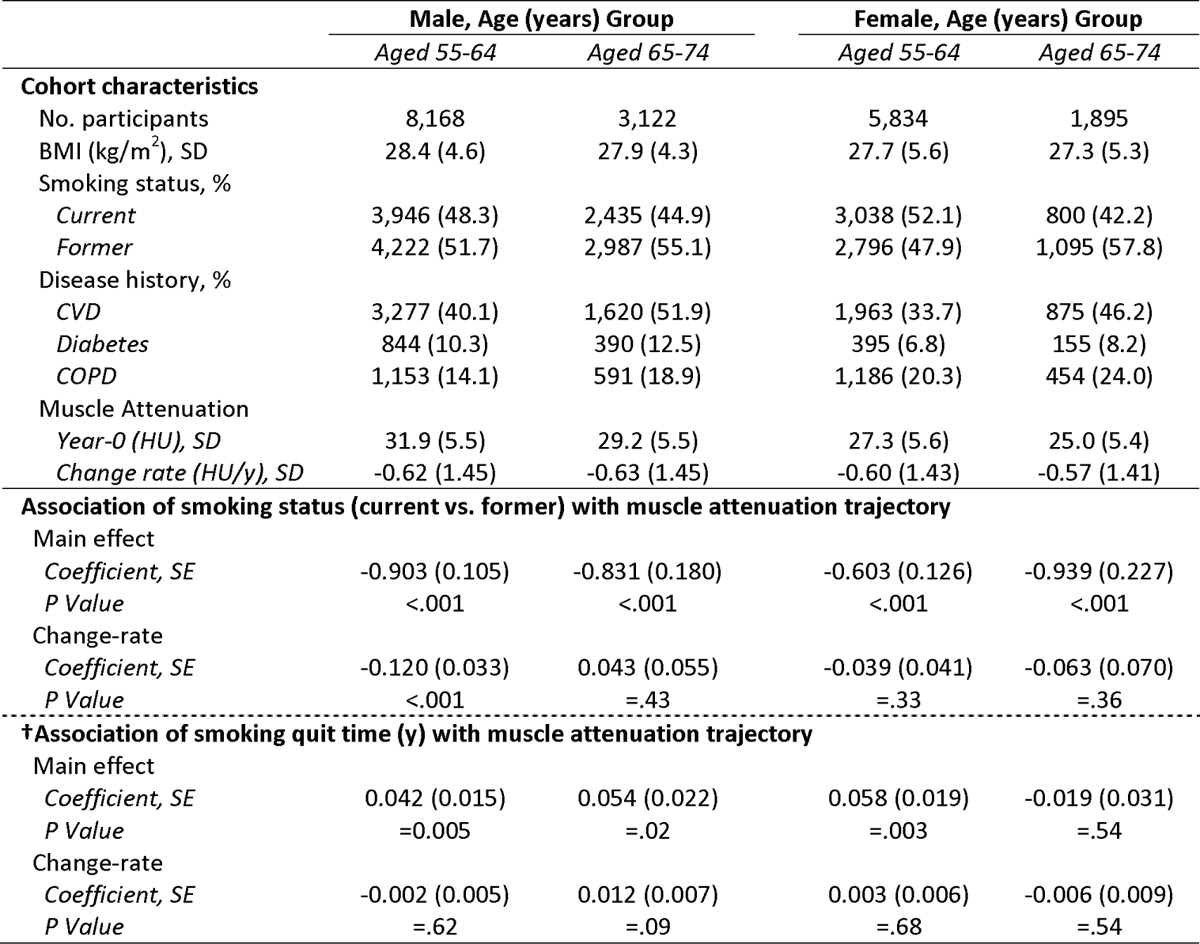
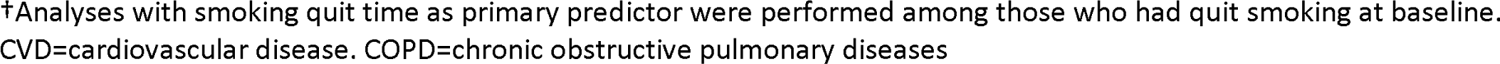
Association of baseline smoking status with longitudinal trajectory of muscle attenuation in analysis stratified by age. The first part shows the characteristics of each subgroup, while the second part shows regression coefficients of fully adjusted linear mix-effects models for each subgroup.

|  | Male, Age (years) Group |  | Female, Age (years) Group |  |
| --- | --- | --- | --- | --- |
|  | Aged 55-64 | Aged 65-74 | Aged 55-64 | Aged 65-74 |
| **Cohort characteristics** |  |  |  |  |
| No. participants | 8,168 | 3,122 | 5,834 | 1,895 |
| BMI (kg/m<sup>2</sup>), SD | 28.4 (4.6) | 27.9 (4.3) | 27.7 (5.6) | 27.3 (5.3) |
| Smoking status, % |  |  |  |  |
| Current | 3,946 (48.3) | 2,435 (44.9) | 3,038 (52.1) | 800 (42.2) |
| Former | 4,222 (51.7) | 2,987 (55.1) | 2,796 (47.9) | 1,095 (57.8) |
| Disease history, % |  |  |  |  |
| CVD | 3,277 (40.1) | 1,620 (51.9) | 1,963 (33.7) | 875 (46.2) |
| Diabetes | 844 (10.3) | 390 (12.5) | 395 (6.8) | 155 (8.2) |
| COPD | 1,153 (14.1) | 591 (18.9) | 1,186 (20.3) | 454 (24.0) |
| Muscle Attenuation |  |  |  |  |
| Year-0 (HU), SD | 31.9 (5.5) | 29.2 (5.5) | 27.3 (5.6) | 25.0 (5.4) |
| Change rate (HU/y), SD | -0.62 (1.45) | -0.63 (1.45) | -0.60 (1.43) | -0.57 (1.41) |
| **Association of smoking status (current vs. former) with muscle attenuation trajectory** |  |  |  |  |
| Main effect |  |  |  |  |
| Coefficient, SE | -0.903 (0.105) | -0.831 (0.180) | -0.603 (0.126) | -0.939 (0.227) |
| P Value | <.001 | <.001 | <.001 | <.001 |
| Change-rate |  |  |  |  |
| Coefficient, SE | -0.120 (0.033) | 0.043 (0.055) | -0.039 (0.041) | -0.063 (0.070) |
| P Value | <.001 | =.43 | =.33 | =.36 |
| **†Association of smoking quit time (y) with muscle attenuation trajectory** |  |  |  |  |
| Main effect |  |  |  |  |
| Coefficient, SE | 0.042 (0.015) | 0.054 (0.022) | 0.058 (0.019) | -0.019 (0.031) |
| P Value | =0.005 | =.02 | =.003 | =.54 |
| Change-rate |  |  |  |  |
| Coefficient, SE | -0.002 (0.005) | 0.012 (0.007) | 0.003 (0.006) | -0.006 (0.009) |
| P Value | =.62 | =.09 | =.68 | =.54 |

†Analyses with smoking quit time as primary predictor were performed among those who had quit smoking at baseline. CVD=cardiovascular disease. COPD=chronic obstructive pulmonary diseases

## 4. Discussion

The primary goal of this secondary analysis of the National Lung Screening Trial was to assess whether smoking cessation has an impact on the progression of skeletal muscle fat infiltration with aging within the lung cancer screening population. Utilizing a previously validated artificial intelligence algorithm, we characterized the longitudinal trajectory of skeletal muscle attenuation by analyzing annual lung screening low-dose CT scans of the chest over a two-year observational period. Here, a lower attenuation indicates increased adipose (fat) infiltration. After adjusting for confounding factors such as body mass index, pack-years, and percent emphysema, active smoking predicted a reduced baseline attenuation in both males and females, and a swifter decline in attenuation in males but not females. Furthermore, among individuals who had quit smoking, a longer duration of smoking cessation was directly associated with improved skeletal muscle quality, as reflected by increased attenuation, in both males and females. The relationship between smoking status and the trajectory of muscle attenuation were influenced by sex, age, and obesity. Notably, the associations between active smoking and decreased muscle attenuation were accentuated with higher levels of obesity. Additionally, the accelerated decline in attenuation associated with active smoking were more prominent at younger age, particularly in females. These findings underscore the important role of smoking cessation in preserving muscle health for older individuals with a significant smoking history.

Muscle attenuation derived from lung screening LDCT could serve as a reliable and reproducible metric for assessing skeletal muscle quality for lung cancer screening participants. Previous research has established a negative correlation between muscle attenuation and muscle lipid concentration (23), as well as a positive association with muscle strength (22). Support for this was also found in *in vivo* animal studies, which confirmed the causative relationship between increased muscle adiposity and decreased muscle function (46). The examination of skeletal muscles through the analysis of multiple axial images as undertaken in the present study covered diverse muscle groups across the fifth, eight, and 10th vertebral levels, enabling a comprehensive encoding of anatomic information linked to skeletal muscle function (47–49). The utilization of fully automatic AI-based solutions enhances the feasibility of deploying such assessments at population scale (27, 30, 33, 34). Previous investigations based on the NLST data have affirmed the added prognostic value of AI measurements of the muscle attenuation, particularly in mortality risk prediction (27, 30). This current study extends these findings by showcasing the potential utility of AI-based muscle attenuation in monitoring the longitudinal trajectory of skeletal muscle health in this population.

Smoking-induced muscle composition alternations are closely relevant to the metabolic perturbations (4, 8, 50, 51). Increased insulin resistance is associated with chronic cigarette smoking, which can possibly be explained by nicotine-induced activation of mammalian target of rapamycin in skeletal muscle (52, 53). Free fatty acid oversupply in plasma results directly from insulin resistance-induced excessive adipocyte lipolysis and is believed to be the primary source of lipid accumulation in skeletal muscle (54). Elevated levels of lipid-derived metabolites, coupled with excess free fatty acid, can further compromise skeletal muscle insulin signaling, trigger pro-inflammatory pathways, and disrupt the balance between muscle protein degradation and synthesis (1, 54). In a recent study utilizing data from the Coronary Artery Risk Development in Young Adults (CARDIA) cohort (32), middle-aged (aged 42-58 years) participants who had ever smoked exhibited lower abdominal muscle attenuation and higher intermuscular adipose tissue volume compared to those who had never smoked. Similarly, findings from the Age, Gene/Environment Susceptibility-Reykjavik (AGES-Reykjavik) study (31) indicated that older participants (aged 66-95 years) with a history of smoking displayed reduced CT-derived quadriceps muscle attenuation in comparison to those who had never smoked. In combination, these studies affirm the connection between cigarette smoking and elevated fatty infiltration in skeletal muscle, observed across middle-aged and older populations. However, the potential benefits of smoking cessation among those who ever smoked were not observed or explicitly investigated in this context. Both evidence from animal models and findings from small-scale clinical studies indicate that the direct adverse effects of smoking on insulin sensitivity and skeletal muscle function are likely reversible, even within relatively short period following smoking cessation (4, 8, 55, 56). Interestingly, in unadjusted analysis, we noted a paradoxical finding of even more severe muscle fat infiltration (indicated by lowered attenuation) among individuals who had quit smoking. This observation can be explained by the observed higher BMI and advanced age, both of which are positively linked to muscle fat infiltration (16, 57), among those who ceased smoking. Additionally, individuals who had quit smoking exhibited increased diagnosis history of comorbidities associated with smoking, such as cardiovascular diseases, diabetes, and increased emphysema. Indeed, after accounting for these confounding factors, active smoking predicted a lowered baseline muscle attenuation in both males and females. Our analyses with incrementally added confounding factors further revealed the particular importance of adjusting for BMI to unveil previously obscured associations. In BMI-stratified analyses, we observed a growing trend of the strength of associations between smoking status (current vs. former) and baseline muscle attenuation in both males and females.

This suggests that the reduction in fatty infiltration following smoking cessation could be amplified in the presence of higher baseline obesity. This aligns with the important roles of insulin resistance and obesity in the mechanisms of excessive skeletal muscle fat infiltration.

While the onset of frailty is substantially associated with older age (e.g., > 65 years), the accelerated decline of muscle strength begins in midlife (12). In our study, we affirmed a population-wise decline in muscle quality in both younger (aged 55-64 years) and older (aged 65-74 years) age groups, as indicated by the relatively consistent muscle attenuation decline rates across the two age groups in both males and females. In age-stratified analyses, notably, our data suggest that the effects of smoking cessation on decelerating muscle quality decline were predominant at younger age (aged 55-64 years) in males and may happened at an even earlier age (< 55 years) in females. First, we observed an accelerated decline associated with active smoking in the follow-up period specifically in younger males, but not in older males or females within either age group. Second, among individuals who had quit smoking, there was an association between longer smoking cessation duration and increased attenuation in both age groups for males and in younger females, yet this association was not observed in older females. In consistent with our findings, a longitudinal research based on the Health, Aging, and Body Composition Study led to the conclusion that the accelerated aging-related decline in physical function associated with obstructive lung disease and smoking among older adults (aged 74±3 years) was likely occurred at a younger age (58). Similarly, AGES-Reykjavik study found no apparent associations between duration since smoking cessation and quadriceps muscle attenuation among the older aged participants (aged 76.4 ± 5.5 years), while an association between a former smoking history and decreased muscle attenuation was observed (31). In combination, these findings suggest the existence of smoking-induced irreversible, residual effects that were accumulated at early age. This could be explained by the smoking-induced irreversible damage to the lung function, as well as the progressive fibrosis and lipid deposition accompanied with prolonged process of muscle damage and regeneration, which further inhibit the regenerative capability of skeletal muscle (5, 59, 60).

Lung cancer screening serves as a “teachable moment” for smoking cessation (19–21), which also forms an opportunity for preventive targeted interventions for individuals at risks of developing frailty. Our findings indicate that, for older individuals with a heavy smoking history, quitting smoking, particularly at middle age, can potentially help to preserve muscle health with the promise of decreasing or postponing the onset of frailty at older age. Meanwhile, the imaging evidence of accelerated muscle degradation observed in annual lung screening LDCT may provide an important extra stimulus to further support the cessation efforts.

### Strengths and Limitations

Unlike previous cross-sectional studies, our primary focus was to evaluate the association between smoking status and longitudinal trajectory in skeletal muscle fat infiltration assessed on CT. Utilizing a previously validated AI-based measurement solution, we presented and analyzed the longitudinal muscle attenuation data for a large-scale lung cancer screening cohort. However, this study has multiple limitations. We employed linear mixed-effects models to assess the associations between baseline smoking status and muscle attenuation trajectory, which can account for the intra-subject measurement correlations in longitudinal repeated measurements. However, the current study design cannot incorporate the possible smoking status change in the observational period. In the present study, to adjust for the confounding factors potentially associated with skeletal muscle fat infiltration, we included multiple factors available in NLST tabular data including BMI, demographic information, disease history, as well as quantitative emphysema measured on CT. However, it is known that lifestyle factors other than smoking, such as the food intake, routine physical activity level (32), can impact metabolic status and influence muscle adiposity. Moreover, smoking cessation could be a part of a systematic adaptation to a healthier lifestyle. Due to lack of relevant data elements, we were unable to account for these lifestyle factors in our models. Our study was secondary analysis based on the NLST data, which may limit the generalizability of the conclusion to another population. Finally, this is a single arm observational study. Therefore, the observed associations do not deduce causation.

## Conclusions

In this study, we investigated the impact of smoking cessation on the age-related progression of skeletal muscle fat infiltration among older individuals with a heavy smoking history who participated NLST. Our findings indicate that active smoking not only associated with increased baseline muscle fat infiltration but also induced an accelerated progression of fatty infiltration particularly in males. Age-stratified analyses unveiled that smoking-associated acceleration of fatty infiltration was likely happened at younger age, which could explain the unobserved acceleration in females. Future studies are needed to investigate smoking-associated impact on the trajectory of muscle fat infiltration in population with a broader age range. Nevertheless, these findings suggest smoking cessation, among its myriad other health benefits, may also promote skeletal muscle health among high-risk individuals undergoing lung cancer screening.

## Data Availability

Data analyzed during the study were provided by a third party. Requests for data should be directed to the provider indicated in the Acknowledgments.

https://cdas.cancer.gov/nlst/

## Acknowledgement

The authors thank the National Cancer Institute (NCI) for access to NCI’s data collected by the National Lung Screening Trial. The statements contained herein are solely those of the authors and do not represent or imply concurrence or endorsement by NCI.

## References

1. Jaitovich A, Barreiro E. Skeletal Muscle Dysfunction in Chronic Obstructive Pulmonary Disease. What We Know and Can Do for Our Patients. American Journal of Respiratory and Critical Care Medicine 2018; 198: 175–186.

2. Barnes PJ, Celli BR. Systemic manifestations and comorbidities of COPD. European Respiratory Journal 2009; 33: 1165–1185.

3. Maltais F, Decramer M, Casaburi R, Barreiro E, Burelle Y, Debigaré R, Dekhuijzen PNR, Franssen F, Gayan-Ramirez G, Gea J, Gosker HR, Gosselink R, Hayot M, Hussain SNA, Janssens W, Polkey MI, Roca J, Saey D, Schols AMWJ, Spruit MA, Steiner M, Taivassalo T, Troosters T, Vogiatzis I, Wagner PD. An Official American Thoracic Society/European Respiratory Society Statement: Update on Limb Muscle Dysfunction in Chronic Obstructive Pulmonary Disease. American Journal of Respiratory and Critical Care Medicine 2014; 189: e15–e62.

4. Degens H, Gayan-Ramirez G, van Hees HWH. Smoking-induced Skeletal Muscle Dysfunction. From Evidence to Mechanisms. American Journal of Respiratory and Critical Care Medicine 2015; 191: 620–625.

5. Barreiro E, Sznajder JI, Nader GA, Budinger GRS. Muscle Dysfunction in Patients with Lung Diseases. A Growing Epidemic. American Journal of Respiratory and Critical Care Medicine 2015; 191: 616–619.

6. Barreiro E, Peinado VI, Galdiz JB, Ferrer E, Marin-Corral J, Sánchez F, Gea J, Barberà JA. Cigarette Smoke–induced Oxidative Stress: A role in chronic obstructive pulmonary disease skeletal muscle dysfunction. American Journal of Respiratory and Critical Care Medicine 2010; 182: 477–488.

7. Montes de Oca M, Loeb E, Torres SH, De Sanctis J, Hernández N, Tálamo C. Peripheral Muscle Alterations in Non-COPD Smokers. Chest 2008; 133: 13–18.

8. Chan SMH, Cerni C, Passey S, Seow HJ, Bernardo I, van der Poel C, Dobric A, Brassington K, Selemidis S, Bozinovski S, Vlahos R. Cigarette Smoking Exacerbates Skeletal Muscle Injury without Compromising Its Regenerative Capacity. American Journal of Respiratory Cell and Molecular Biology 2020; 62: 217–230.

9. Krist AH, Davidson KW, Mangione CM, Barry MJ, Cabana M, Caughey AB, Davis EM, Donahue KE, Doubeni CA, Kubik M, Landefeld CS, Li L, Ogedegbe G, Owens DK, Pbert L, Silverstein M, Stevermer J, Tseng C-W, Wong JB. Screening for Lung Cancer. Jama 2021; 325.

10. Kojima G, Iliffe S, Jivraj S, Liljas A, Walters K. Does current smoking predict future frailty? The English longitudinal study of ageing. Age and Ageing 2018; 47: 126–131.

11. Kojima G, Iliffe S, Walters K. Smoking as a predictor of frailty: a systematic review. BMC Geriatrics 2015; 15.

12. Xue Q-L. The Frailty Syndrome: Definition and Natural History. Clinics in Geriatric Medicine 2011; 27: 1–15.

13. Miljkovic I, Zmuda JM. Epidemiology of myosteatosis. Current Opinion in Clinical Nutrition and Metabolic Care 2010; 13: 260–264.

14. Correa-de-Araujo R, Addison O, Miljkovic I, Goodpaster BH, Bergman BC, Clark RV, Elena JW, Esser KA, Ferrucci L, Harris-Love MO, Kritchevsky SB, Lorbergs A, Shepherd JA, Shulman GI, Rosen CJ. Myosteatosis in the Context of Skeletal Muscle Function Deficit: An Interdisciplinary Workshop at the National Institute on Aging. Frontiers in Physiology 2020; 11.

15. Addison O, Marcus RL, LaStayo PC, Ryan AS. Intermuscular Fat: A Review of the Consequences and Causes. International Journal of Endocrinology 2014; 2014: 1–11.

16. Goodpaster BH, Theriault R, Watkins SC, Kelley DE. Intramuscular lipid content is increased in obesity and decreased by weight loss. Metabolism 2000; 49: 467–472.

17. Hilton TN, Tuttle LJ, Bohnert KL, Mueller MJ, Sinacore DR. Excessive Adipose Tissue Infiltration in Skeletal Muscle in Individuals With Obesity, Diabetes Mellitus, and Peripheral Neuropathy: Association With Performance and Function. Physical Therapy 2008; 88: 1336–1344.

18. Bianchi L, Volpato S. Muscle dysfunction in type 2 diabetes: a major threat to patient’s mobility and independence. Acta Diabetologica 2016; 53: 879–889.

19. Tammemägi MC, Berg CD, Riley TL, Cunningham CR, Taylor KL. Impact of Lung Cancer Screening Results on Smoking Cessation. JNCI: Journal of the National Cancer Institute 2014; 106.

20. Taylor KL, Cox LS, Zincke N, Mehta L, McGuire C, Gelmann E. Lung cancer screening as a teachable moment for smoking cessation. Lung Cancer 2007; 56: 125–134.

21. Wiener RS, Gould MK, Arenberg DA, Au DH, Fennig K, Lamb CR, Mazzone PJ, Midthun DE, Napoli M, Ost DE, Powell CA, Rivera MP, Slatore CG, Tanner NT, Vachani A, Wisnivesky JP, Yoon SH. An Official American Thoracic Society/American College of Chest Physicians Policy Statement: Implementation of Low-Dose Computed Tomography Lung Cancer Screening Programs in Clinical Practice. American Journal of Respiratory and Critical Care Medicine 2015; 192: 881–891.

22. Goodpaster BH, Carlson CL, Visser M, Kelley DE, Scherzinger A, Harris TB, Stamm E, Newman AB. Attenuation of skeletal muscle and strength in the elderly: The Health ABC Study. Journal of Applied Physiology 2001; 90: 2157–2165.

23. Goodpaster BH, Kelley DE, Thaete FL, He J, Ross R. Skeletal muscle attenuation determined by computed tomography is associated with skeletal muscle lipid content. Journal of Applied Physiology 2000; 89: 104–110.

24. Mühlberg A, Museyko O, Bousson V, Pottecher P, Laredo J-D, Engelke K. Three-dimensional Distribution of Muscle and Adipose Tissue of the Thigh at CT: Association with Acute Hip Fracture. Radiology 2019; 290: 426–434.

25. Visser M, Goodpaster BH, Kritchevsky SB, Newman AB, Nevitt M, Rubin SM, Simonsick EM, Harris TB. Muscle Mass, Muscle Strength, and Muscle Fat Infiltration as Predictors of Incident Mobility Limitations in Well-Functioning Older Persons. The Journals of Gerontology Series A: Biological Sciences and Medical Sciences 2005; 60: 324–333.

26. Tattersall MC, Lee KE, Tsuchiya N, Osman F, Korcarz CE, Hansen KM, Peters MC, Fahy JV, Longhurst CA, Dunican E, Wentzel SE, Leader JK, Israel E, Levy BD, Castro M, Erzurum SC, Lempel J, Moore WC, Bleecker ER, Phillips BR, Mauger DT, Hoffman EA, Fain SB, Reeder SB, Sorkness RL, Jarjour NN, Denlinger LC, Schiebler ML. Skeletal Muscle Adiposity and Lung Function Trajectory in the Severe Asthma Research Program. American Journal of Respiratory and Critical Care Medicine 2023; 207: 475–484.

27. Xu K, Khan MS, Li TZ, Gao R, Terry JG, Huo Y, Lasko TA, Carr JJ, Maldonado F, Landman BA, Sandler KL. AI Body Composition in Lung Cancer Screening - Added Value Beyond Lung Cancer Detection. Radiology 2023; 60.

28. Miljkovic I, Kuipers AL, Cauley JA, Prasad T, Lee CG, Ensrud KE, Cawthon PM, Hoffman AR, Dam T-T, Gordon CL, Zmuda JM. Greater Skeletal Muscle Fat Infiltration Is Associated With Higher All-Cause and Cardiovascular Mortality in Older Men. The Journals of Gerontology Series A: Biological Sciences and Medical Sciences 2015; 70: 1133–1140.

29. Larsen B, Bellettiere J, Allison M, McClelland RL, Miljkovic I, Vella CA, Ouyang P, De-guzman KR, Criqui M, Unkart J. Muscle area and density and risk of all-cause mortality: The Multi-Ethnic Study of Atherosclerosis. Metabolism 2020; 111.

30. Lenchik L, Barnard R, Boutin RD, Kritchevsky SB, Chen H, Tan J, Cawthon PM, Weaver AA, Hsu F-C, Melzer D. Automated Muscle Measurement on Chest CT Predicts All-Cause Mortality in Older Adults From the National Lung Screening Trial. The Journals of Gerontology: Series A 2021; 76: 277–285.

31. Marques EA, Elbejjani M, Frank-Wilson AW, Gudnason V, Sigurdsson G, Lang TF, Jonsson PV, Sigurdsson S, Aspelund T, Siggeirsdottir K, Launer L, Eiriksdottir G, Harris TB. Cigarette Smoking Is Associated With Lower Quadriceps Cross-sectional Area and Attenuation in Older Adults. Nicotine & Tobacco Research 2020; 22: 935–941.

32. Terry JG, Hartley KG, Steffen LM, Nair S, Alman AC, Wellons MF, Jacobs DR, Tindle HA, Carr JJ. Association of smoking with abdominal adipose deposition and muscle composition in Coronary Artery Risk Development in Young Adults (CARDIA) participants at mid-life: A population-based cohort study. PLOS Medicine 2020; 17.

33. Xu K, Gao R, Tang Y, Deppen S, Sandler K, Kammer M, Antic S, Maldonado F, Huo Y, Khan M, Landman BA, Išgum I, Colliot O. Extending the value of routine lung screening CT with quantitative body composition assessment. Medical Imaging 2022: Image Processing; 2022.

34. Xu K, Li T, Khan MS, Gao R, Antic SL, Huo Y, Sandler KL, Maldonado F, Landman BA. Body composition assessment with limited field-of-view computed tomography: A semantic image extension perspective. Medical Image Analysis 2023: 102852.

35. National Lung Screening Trial Research Team. Reduced Lung-Cancer Mortality with Low-Dose Computed Tomographic Screening. N Engl J Med 2011; 365: 395–409.

36. National Lung Screening Trial Research Team, Aberle DR, Berg CD, Black WC, Church TR, Fagerstrom RM, Galen B, Gareen IF, Gatsonis C, Goldin J, Gohagan JK, Hillman B, Jaffe C, Kramer BS, Lynch D, Marcus PM, Schnall M, Sullivan DC, Sullivan D, Zylak CJ. The National Lung Screening Trial: Overview and Study Design. Radiology 2011; 258: 243–253.

37. Cagnon CH, Cody DD, McNitt-Gray MF, Seibert JA, Judy PF, Aberle DR. Description and Implementation of a Quality Control Program in an Imaging-Based Clinical Trial. Acad Radiol 2006; 13: 1431–1441.

38. Gierada DS, Garg K, Nath H, Strollo DC, Fagerstrom RM, Ford MB. CT Quality Assurance in the Lung Screening Study Component of the National Lung Screening Trial: Implications for Multicenter Imaging Trials. AJR Am J Roentgenol 2009; 193: 419–424.

39. Matelsky J, Kiar G, Johnson E, Rivera C, Toma M, Gray-Roncal W. Container-Based Clinical Solutions for Portable and Reproducible Image Analysis. Journal of Digital Imaging 2018; 31: 315–320.

40. Hofmanninger J, Prayer F, Pan J, Röhrich S, Prosch H, Langs G. Automatic lung segmentation in routine imaging is primarily a data diversity problem, not a methodology problem. European Radiology Experimental 2020; 4.

41. Oelsner EC, Carr JJ, Enright PL, Hoffman EA, Folsom AR, Kawut SM, Kronmal RA, Lederer DJ, Lima JAC, Lovasi GS, Smith BM, Shea SJ, Barr RG. Per cent emphysema is associated with respiratory and lung cancer mortality in the general population: a cohort study. Thorax 2016; 71: 624–632.

42. Hoffman EA, Ahmed FS, Baumhauer H, Budoff M, Carr JJ, Kronmal R, Reddy S, Barr RG. Variation in the Percent of Emphysema-like Lung in a Healthy, Nonsmoking Multiethnic Sample. The MESA Lung Study. Annals of the American Thoracic Society 2014; 11: 898–907.

43. Fitzmaurice GM, Ravichandran C. A Primer in Longitudinal Data Analysis. Circulation 2008; 118: 2005–2010.

44. Baraghoshi D, Strand M, Humphries SM, San José Estépar R, Vegas Sanchez-Ferrero G, Charbonnier J-P, Latisenko R, Silverman EK, Crapo JD, Lynch DA. Quantitative CT Evaluation of Emphysema Progression over 10 Years in the COPDGene Study. Radiology 2023; 307.

45. Wang M, Aaron CP, Madrigano J, Hoffman EA, Angelini E, Yang J, Laine A, Vetterli TM, Kinney PL, Sampson PD, Sheppard LE, Szpiro AA, Adar SD, Kirwa K, Smith B, Lederer DJ, Diez-Roux AV, Vedal S, Kaufman JD, Barr RG. Association Between Long-term Exposure to Ambient Air Pollution and Change in Quantitatively Assessed Emphysema and Lung Function. Jama 2019; 322.

46. Biltz NK, Collins KH, Shen KC, Schwartz K, Harris CA, Meyer GA. Infiltration of intramuscular adipose tissue impairs skeletal muscle contraction. The Journal of Physiology 2020; 598: 2669–2683.

47. Troschel AS, Troschel FM, Best TD, Gaissert HA, Torriani M, Muniappan A, Van Seventer EE, Nipp RD, Roeland EJ, Temel JS, Fintelmann FJ. Computed Tomography–based Body Composition Analysis and Its Role in Lung Cancer Care. Journal of Thoracic Imaging 2020; 35: 91–100.

48. Fintelmann FJ. Body Composition Analysis on Chest CT Scans: A Value Proposition for Lung Cancer Care. Radiology 2023; 308.

49. Best TD, Mercaldo SF, Bryan DS, Marquardt JP, Wrobel MM, Bridge CP, Troschel FM, Javidan C, Chung JH, Muniappan A, Bhalla S, Meyers BF, Ferguson MK, Gaissert HA, Fintelmann FJ. Multilevel Body Composition Analysis on Chest Computed Tomography Predicts Hospital Length of Stay and Complications After Lobectomy for Lung Cancer. Annals of Surgery 2022; 275: e708–e715.

50. Caron M-A, Morissette MC, Thériault M-E, Nikota JK, Stämpfli MR, Debigaré R. Alterations in Skeletal Muscle Cell Homeostasis in a Mouse Model of Cigarette Smoke Exposure. PLoS ONE 2013; 8.

51. Darabseh MZ, Maden-Wilkinson TM, Welbourne G, Wüst RCI, Ahmed N, Aushah H, Selfe J, Morse CI, Degens H. Fourteen days of smoking cessation improves muscle fatigue resistance and reverses markers of systemic inflammation. Scientific Reports 2021; 11.

52. Bergman BC, Perreault L, Hunerdosse D, Kerege A, Playdon M, Samek AM, Eckel RH. Novel and Reversible Mechanisms of Smoking-Induced Insulin Resistance in Humans. Diabetes 2012; 61: 3156–3166.

53. Facchini FS, Hollenbeck CB, Jeppesen J, Ida Chen YD, Reaven GM. Insulin resistance and cigarette smoking. The Lancet 1992; 339: 1128–1130.

54. Eckardt K, Taube A, Eckel J. Obesity-associated insulin resistance in skeletal muscle: Role of lipid accumulation and physical inactivity. Reviews in Endocrine and Metabolic Disorders 2011; 12: 163–172.

55. Thatcher TH, Caron M-A, Morissette MC, Thériault M-E, Nikota JK, Stämpfli MR, Debigaré R. Alterations in Skeletal Muscle Cell Homeostasis in a Mouse Model of Cigarette Smoke Exposure. PLoS ONE 2013; 8.

56. Seet RCS, Loke WM, Khoo CM, Chew SE, Chong WL, Quek AML, Lim ECH, Halliwell B. Acute effects of cigarette smoking on insulin resistance and arterial stiffness in young adults. Atherosclerosis 2012; 224: 195–200.

57. Aubrey J, Esfandiari N, Baracos VE, Buteau FA, Frenette J, Putman CT, Mazurak VC. Measurement of skeletal muscle radiation attenuation and basis of its biological variation. Acta Physiologica 2014; 210: 489–497.

58. van den Borst B, Koster A, Yu B, Gosker HR, Meibohm B, Bauer DC, Kritchevsky SB, Liu Y, Newman AB, Harris TB, Schols AMWJ. Is age-related decline in lean mass and physical function accelerated by obstructive lung disease or smoking? Thorax 2011; 66: 961–969.

59. Sciorati C, Clementi E, Manfredi AA, Rovere-Querini P. Fat deposition and accumulation in the damaged and inflamed skeletal muscle: cellular and molecular players. Cellular and Molecular Life Sciences 2015; 72: 2135–2156.

60. Mahdy MAA. Skeletal muscle fibrosis: an overview. Cell and Tissue Research 2018; 375: 575–588.

